# Durability of Omicron-neutralizing serum activity following mRNA booster immunization in elderly individuals

**DOI:** 10.1101/2022.02.02.22270302

**Authors:** Kanika Vanshylla, Pinkus Tober-Lau, Henning Gruell, Friederike Münn, Ralf Eggeling, Nico Pfeifer, N. Han Le, Irmgard Landgraf, Florian Kurth, Leif E. Sander, Florian Klein

**Author notes:** These authors contributed equally (Kanika Vanshylla, Pinkus Tober-Lau, Henning Gruell, Florian Kurth, Leif E. Sander, Florian Klein).

## Abstract

Elderly individuals are at high risk for severe COVID-19. Due to modest vaccine responses compared to younger individuals and the time elapsed since prioritized vaccinations, the emerging immune-evasive Omicron variant of SARS-CoV-2 is a particular concern for the elderly. Here we longitudinally determined SARS-CoV-2-neutralizing serum activity against different variants in a cohort of 37 individuals with a median age of 82 years. Participants were followed for 10 months after an initial two-dose BNT162b2 vaccination and up to 4.5 months after a BNT162b2 booster. Detectable Omicron-neutralizing activity was nearly absent after two vaccinations but elicited in 89% of individuals by the booster immunization. Neutralizing titers against the Wu01, Delta, and Omicron variants showed similar post-boost declines and 81% of individuals maintained detectable activity against Omicron. Our study demonstrates the mRNA booster effectiveness in inducing Omicron neutralizing activity and provides critical information on vaccine response durability in the highly vulnerable elderly population.

## Text

Advanced age is a critical risk factor for morbidity and mortality associated with SARS-CoV-2 infection. Elderly individuals have therefore been prioritized for COVID-19 vaccination. Moreover, lower vaccine immunogenicity in older compared to younger individuals and more pronounced waning of humoral immunity have prompted early booster campaigns in the elderly.^1^ The recently emerged Omicron variant (BA.1) of SARS-CoV-2 is associated with a marked reduction in sensitivity to vaccine-induced serum neutralizing activity and is therefore of particular concern for the elderly.^2^ However, while booster immunizations can elicit Omicron-neutralizing activity,^3^ their immediate and long-term effects in highly aged individuals are not yet determined. This limits informed guidance of vaccination strategies in this vulnerable population.

Here, we longitudinally determined SARS-CoV-2-neutralizing serum activity in a prospective cohort of 37 individuals with a median age of 82 years (range 76-96 years; appendix p 2).^4^ Participants were followed for 10 months after an initial two-dose BNT162b2 vaccination and up to 4.5 months after a single booster dose of BNT162b2. 50% geometric mean inhibitory serum dilutions (GeoMean ID_50_s) against the Wu01 vaccine strain as well as the Delta and Omicron variants were determined using a pseudovirus assay (appendix p 4).

Following completion of the initial two-dose regimen, sera were collected at 1 (median 26 days, interquartile range [IQR] 25-27; *V1*) and 5 months (median 153 days; IQR 151-154; *V2*). Two doses of BNT162b2 induced detectable neutralizing activity against Wu01 and Delta in the majority of individuals (95% and 84%, respectively), while activity against Omicron was not or only minimally detectable (figure). Over the subsequent 4 months, neutralizing serum titers against Wu01 and Delta declined by 6- (GeoMean ID_50_ of 265 to 42) and 7-fold (GeoMean ID_50_ of 89 to 13), respectively.

**Figure.**
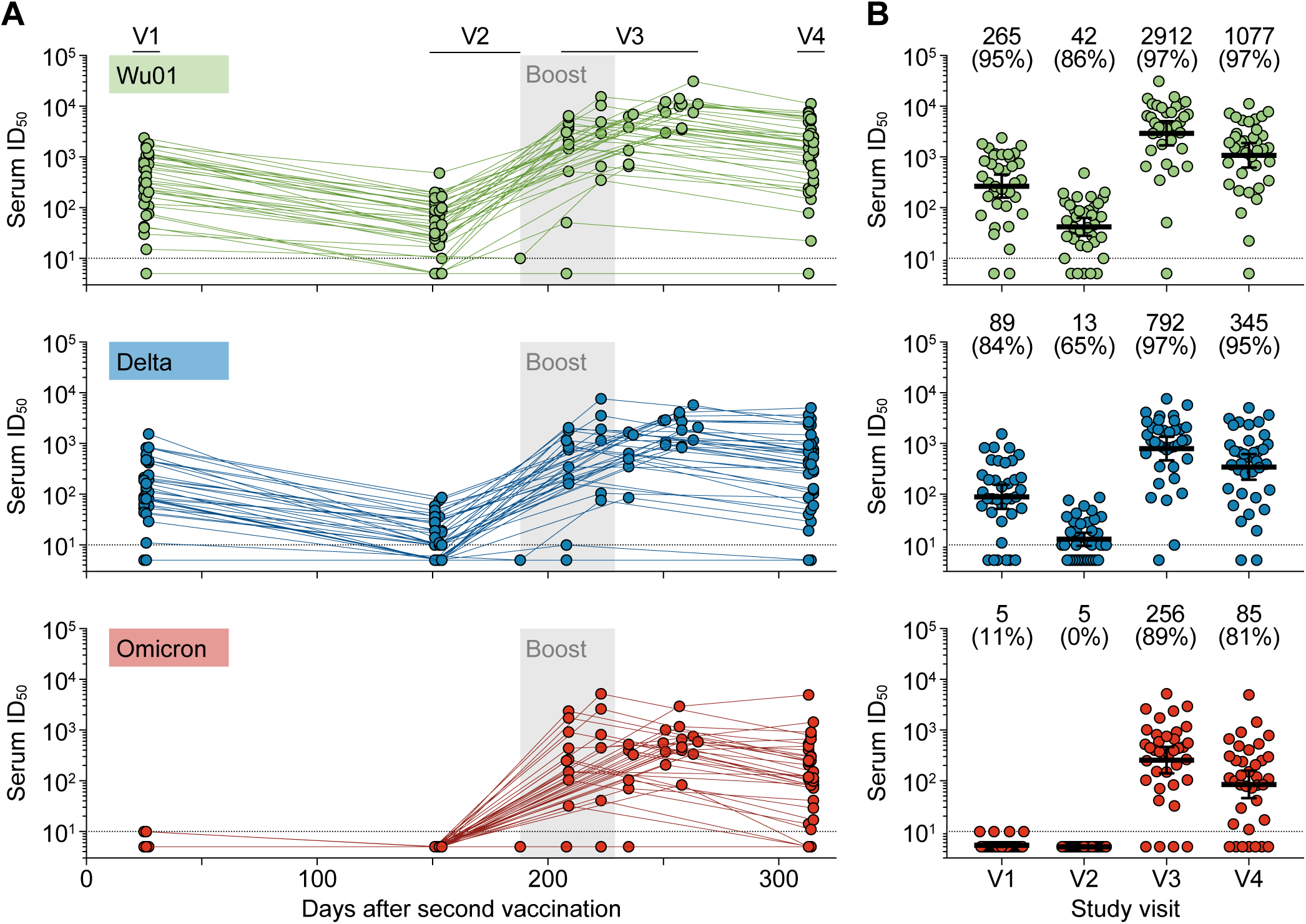
Longitudinal assessment of SARS-CoV-2-neutralizing serum activity in elderly individuals. Serum ID_50_s against the Wu01, Delta, and Omicron variants, determined using pseudovirus neutralization assays. Lines in (A) connect visits for each individual and gray areas indicate booster administration period. Lines in (B) indicate geometric mean ID_50_s with 95% confidence intervals for each study visit. Numbers show geometric mean ID_50_s and percentage of individuals with detectable neutralizing activity (ID_50_>LLOQ) in parentheses. Dotted lines indicate LLOQ. V1-V4 indicate consecutive study visits. ID_50_, 50% inhibitory dilution; LLOQ, lower limit of quantification.

All individuals received a third dose of BNT162b2 at 7 months (median 209 days, IQR 189-228) and early post-boost serum samples were obtained 1 month later (median 23 days, IQR 21-29, *V3*). Booster immunization resulted in a more than 50-fold increase in neutralizing titers against Wu01 and Delta (GeoMean ID_50_s of 2912 and 750, respectively). Importantly, the booster dose of BNT162b2 elicited a robust neutralizing activity against the Omicron variant (GeoMean ID_50_ of 256) in 33 out of the 37 elderly participants (89%; figure). Of the four non-responders, one participant suffered from an active hematological malignancy and did not develop neutralizing reactivity against all of the tested variants at any time point.

To determine the durability of SARS-CoV-2 serum neutralizing activity after booster immunization in elderly individuals, we obtained follow-up samples 3·5 months (median 106 days, IQR 86-125) after the third vaccination. Neutralizing titers declined by 2·7-, 2·3-, and 3·0-fold to GeoMean ID_50_s of 1077, 345, and 85 against the Wu01, Delta, and Omicron variants, respectively. However, most participants maintained detectable neutralization against Wu01 (97%), Delta (95%), and Omicron (81% total; 91% of individuals with detectable activity at the early post-boost visit V3). To evaluate the rate of the decline in serum neutralizing activity, we used linear mixed-effects models to separately analyze the periods before and after the booster immunization (appendix p 3). Neutralizing activity against the different variants showed similar changes after the booster immunization with estimated half-lives of approximately 52 (95% confidence interval [CI], 46-59), 64 (95% CI 52-83), and 41 (95% CI 34-52) days against Wu01, Delta, and Omicron, respectively (appendix p 3).

In the absence of Omicron-specific vaccines, booster immunizations are critical to restore waning vaccine effectiveness and to reduce the risk of severe outcomes.^5^ Our study demonstrates that booster immunizations can effectively elicit Omicron-neutralizing activity in the majority of aged individuals (median age 82 years). While our analysis was limited to two sampling time points with different observational periods before and after the boost, our results suggest that neutralizing activity against different variants is maintained with similar decay rates. Although neutralizing serum activity does not equal protection from infection, the results suggest that previous experience regarding waning humoral immunity can serve as guidance for vaccination strategies against Omicron in the elderly population.

## Data Availability

All data in included in manuscript, figure or supplement.

## Acknowledgements

We thank all study participants for their dedication to our research. We thank the members of the COVIMMUNIZE/COVIM Study Group for sample acquisition and processing (Y. Ahlgrimm, B. Al-Rim, K. Behn, N. Bethke, H. Bias, D. Briesemeister, C. Conrad, V.M. Corman, C. Dang-Heine, S. Dieckmann, D. Frey, J.-A. Gabelich, J. Gerdes, U. Gläser, L. Hasler, E.T. Helbig, A. Hetey, D. Hillus, W.G. Hirst, A. Horn, C. Hülso, S. Jentzsch, C. von Kalle, L. Kegel, A. Krannich, W. Koch, P. Kopankiewicz, P. Kroneberg, L.J. Lippert, M. Lisy, C. Lüttke, P. de Macedo Gomes, B. Maeß, J. Michel, A. Nitsche, A.-M. Ollech, C. Peiser, A. Pioch, C. Pley, K. Pohl, A. Richter, M. Rönnefarth, L. Ruby, C. Rubisch, A. Sanchez Rezza, I. Schellenberger, V. Schenkel, J. Schlesinger, S. Schmidt, G. Schwanitz, T. Schwarz, S. Senaydin, J. Seybold, A.-S. Sinnigen, A. Solarek, A. Stege, S. Steinbrecher, P. Stubbemann, C. Thibeault, D. Treue, and S. Zvorc). This work was supported through grants from COVIM: NaFoUniMedCovid19 (FKZ: 01KX2021) to LES and FKl, the German Center for Infection Research (DZIF) to FKl, the Federal Institute for Drugs and Medical Devices (V-2021.3 / 1503_68403 / 2021–2022) to FKu and LES, the Deutsche Forschungsgemeinschaft (DFG) SFB-TR84 to LES and CRC1310 to FKl, and a donation from Zalando SE to Charité – Universitätsmedizin Berlin. The funders had no role in study design, data collection and analysis, manuscript preparation, or the decision to submit for publication.

