## Supplement for "Durability of Omicron-neutralizing serum activity following mRNA booster immunization in elderly individuals"

**Supplementary table. Study cohort chacteristics.**

| | |
|---|---|
| **Participants** - *n* | 37 |
| **Demographics** | |
| **Age** - median years (range) | 82 (76-96) |
| **Gender** | |
| Female - *n* (%) | 27 (73%) |
| Male - *n* (%) | 10 (27%) |
| **Reported comorbidities** | |
| Cardiovascular disease - *n* (%) | 29 (78%) |
| Diabetes - *n* (%) | 9 (24%) |
| Respiratory disease - *n* (%) | 5 (14%) |
| Solid malignancy - *n* (%) | 4 (11%) |
| Immunodeficiencies - *n* (%) | 4 (11%) |
| Hematologic malignancy - *n* (%) | 1 (3%) |
| **Body mass index** - median (IQR, range) | 23.4 (21.7-25.3; 18.0-29.6) |
| **Vaccination** | |
| **Vaccine administered** | BNT162b2 |
| **Time between first and second dose** | |
| Median days (IQR; range) | 21 (21; 21) |
| **Time between second and third dose** | |
| Median days (IQR; range) | 209 (189-228; 188-229) |
| **Sampling time points** | |
| **Time after second dose** | |
| V1 - median days (IQR; range) | 26 (25-27; 25-27) |
| V2 - median days (IQR; range) | 153 (151-154; 151-188) |
| V3 - median days (IQR; range) | 235 (209-254; 208-265) |
| V4 - median days (IQR; range) | 314 (313-315; 313-315) |

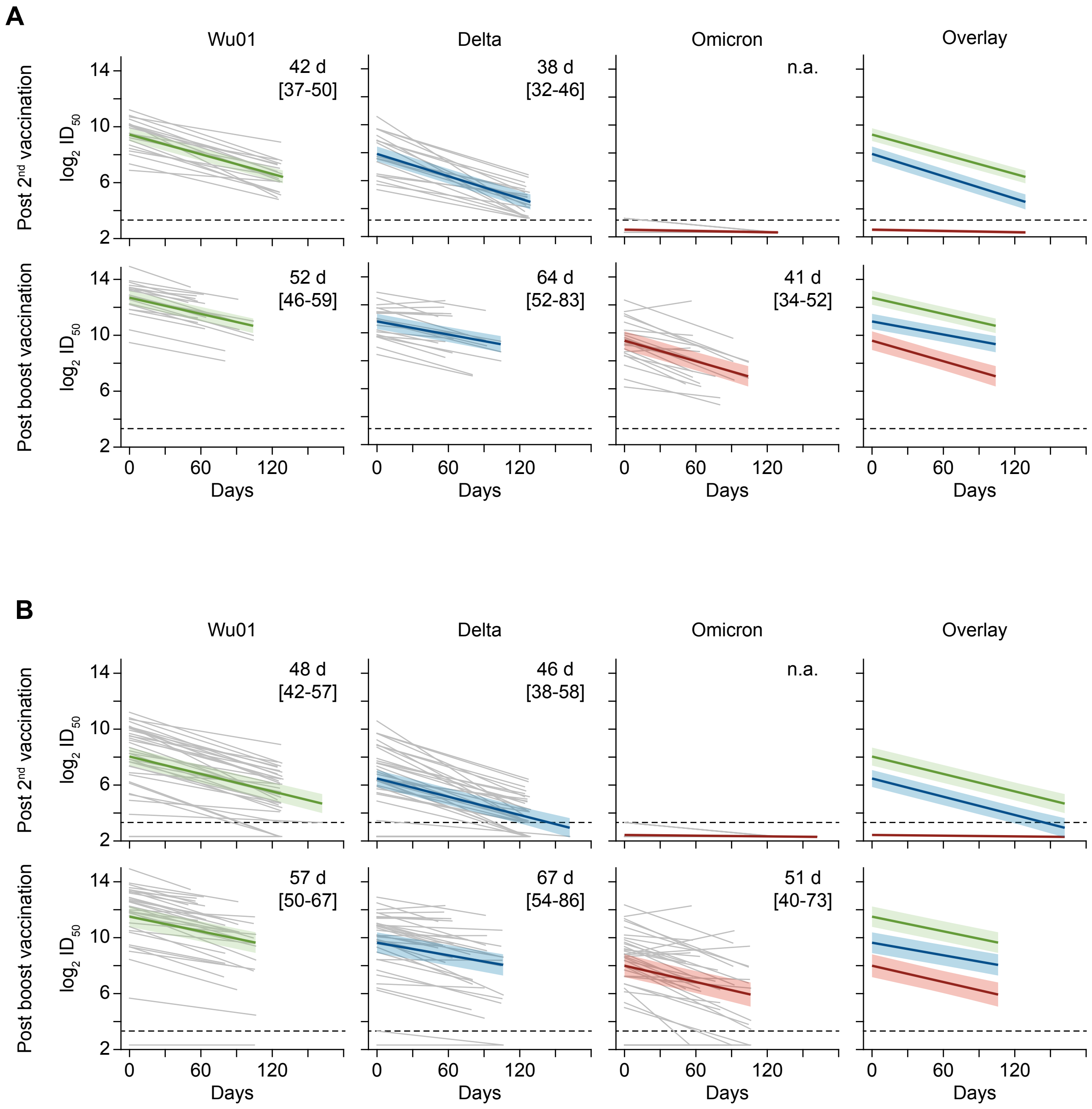

**Supplementary figure. Analysis of declining SARS-CoV-2-neutralizing serum activity in elderly individuals.**
Linear mixed-effects models with common slope and patient-specific intercepts were used to predict the binary logarithm of the serum neutralizing $ID_{50}$. (A) shows the analysis performed with a selection of study participants (*n*=21) with detectable serum neutralizing activity against all variants and time points (excluding the pre-boost period for Omicron), (B) shows the analysis performed with all study participants (*n*=37) . Upper and lower panels show the change in serum neutralizing activity between the V1/V2 and V3/V4 study visits, respectively. Grey lines indicate the change for individual participants. Colored lines indicate the calculated slope for the individual SARS-CoV-2 variant with shaded areas highlighting 95% confidence intervals. Numbers show calculated half-life estimates of serum neutralizing activity and 95% confidence intervals in brackets. Dashed black lines indicate the lower limit of quantification ($ID_{50}$=10). n.a., not applicable.

### Methods

#### Study participants

Elderly participants were enrolled in a longitudinal observational cohort study investigating the immune response to COVID-19 vaccination at a general practice in Berlin, Germany, under the auspices of Charité - Universitätsmedizin Berlin, Germany. Samples were obtained under protocols approved by the ethics committees of Charité - Universitätsmedizin Berlin (EA4/244/20) and of the State of Berlin, as well as by the Federal Institute for Vaccines and Biomedicines (Paul Ehrlich Institute). All study participants provided written informed consent. All serum samples were tested for anti-nucleocapsid antibodies targeting the SARS-CoV-2 nucleocapsid using the microarray-based SeraSpot Anti-SARS-CoV-2 IgG immunoassay (Seramun Diagnostica). Individuals with a history of SARS-CoV-2 infection by anamnesis or laboratory investigation (positive nucleic acid amplification test or anti-nucleocapsid antibodies) were not included in this analysis. Samples obtained at V1 and V2 were included in a previously reported interim analysis of a cohort investigating serum neutralizing activity against the Delta variant.[1,2]

#### Pseudovirus neutralization assays

Serum samples were collected by centrifugation, stored at -80°C until analysis, and heat-inactivated at 56°C for 45 min before use in neutralization assays. Serum neutralizing activity was analyzed using a lentivirus-based SARS-CoV-2 spike pseudovirus system.[3,4] SARS-CoV-2 pseudovirus particles were generated by co-transfection of plasmids encoding HIV-1 Gag/Pol, HIV-1 Tat, HIV-1 Rev, luciferase, and the SARS-CoV-2 spike protein into HEK293T cells using the FuGENE 6 Transfection Reagent (Promega). SARS-CoV-2 spike plasmids encoded for the sequences of the Wu01 spike (EPI_ISL_406716; lacking the cytoplasmic C-terminal 21 amino acids), the Delta variant (B.1.617.2) or the Omicron variant (BA.1 / B.1.1.529.1). Transfected cells were cultured at 37°C and 5% $CO_2$. Virus culture supernatants were harvested between 48 h and 72 h after transfection, filtered using a 0.45 µm filter, and stored at -80°C until use. Serial dilutions of serum (1:3 dilution series starting at 1:10) were co-incubated with pseudovirus supernatants for 1 hour at 37°C and 5% $CO_2$ prior to addition of 293T-ACE2 cells. Following a 48-hour incubation at 37°C and 5% $CO_2$, luciferase activity was determined after addition of luciferin/lysis buffer (10 mM $MgCl_2$, 0.3 mM ATP, 0.5 mM coenzyme A, 17 mM IGEPAL (all Sigma-Aldrich), and 1 mM D-Luciferin (GoldBio) in Tris-HCL) using a microplate reader (Berthold). After subtracting background relative light units

(RLUs) of non-infected cells, 50% serum inhibitory dilutions ($ID_{50}$s) were calculated as the serum dilution resulting in a 50% RLU reduction compared to the untreated virus control wells. $ID_{50}$ values were calculated by using a non-linear fit model to plot an agonist vs. normalized dose response curve with variable slope and least squares fit in GraphPad Prism 7.0. Serum samples not reaching 50% inhibition at the lowest tested dilution of 10 (lower limit of quantification, LLOQ) were assigned a value ½ of the LLOQ ($ID_{50}$=5). Longitudinal samples against each viral variant were tested in parallel within a single assay. All serum samples were tested in duplicates and the average $ID_{50}$s were used for analysis. For samples with a detectable $ID_{50}$ in only one of the duplicates, an $ID_{50}$ equal to the LLOQ ($ID_{50}$=10) was assigned.

**Analysis of SARS-CoV-2-neutralizing serum titer decay**

For analysis of the kinetics of the serum SARS-CoV-2-neutralizing titer, a linear mixed-effects model (R-function lme4::lmer)[5] with common slope and subject-specific intercepts was applied to model the dependence of the $log_2$-transformed $ID_{50}$ on the time difference between the two visits of each observation period (V1/V2, observation period after two vaccinations; V3/V4, observation period after the booster immunization). Half-life estimates were computed as negative inverse of the common slope regression coefficient. 95% confidence intervals were computed using R-function ggeffects::ggpredict.[6] The analysis was performed separately in the full study data set and in the subset of participants showing detectable $ID_{50}$ titers for all viruses at all time points (excluding titers determined against the Omicron variant before booster immunization, based on overall lack of activity in this time period).

**Author contributions**

Conceptualization, KV, PTL, HG, FKu, LES, and FKl; Methodology, KV, PTL, HG, RE, NP, Fku, LES, and FKl; Investigation, KV, PTL, and HG; Resources, PTL, FM, NHL, IL, and L.E.S.; Formal Analysis, KV, HG, RE; Writing - Original Draft, KV, HG, and FKl; Writing – Review & Editing, PTL, RE, NHL, Fku, and LES; Visualization, KV, HG, and FKl; Supervision, NP, FKu, LES, and FKl.; Funding Acquisition, FKu, LES, and FKl.
